# Fentanyl, Heroin, and Methamphetamine-Based Counterfeit Pills Sold at Tourist-Oriented Pharmacies in Mexico: An Ethnographic and Drug Checking Study

**DOI:** 10.1101/2023.01.27.23285123

**Authors:** Joseph Friedman, Morgan Godvin, Caitlin Molina, Ruby Romero, Annick Borquez, Tucker Avra, David Goodman-Meza, Steffanie Strathdee, Philippe Bourgois, Chelsea L. Shover

**Affiliations:** Center for Social Medicine and Humanities, University of California, Los Angeles; The Action Lab, Center for Health Policy and Law, Northeastern University; Division of General Internal Medicine and Health Services Research, University of California, Los Angeles; Division of Infectious Diseases and Global Public Health. Department of Medicine, University of California, San Diego; David Geffen School of Medicine, University of California, Los Angeles; Division of Infectious Diseases, University of California, Los Angeles

## Abstract

**Background:** Fentanyl- and methamphetamine-based counterfeit prescription drugs have driven escalating overdose death rates in the US, however their presence in Mexico has not been assessed. Our ethnographic team has conducted longitudinal research focused on illicit drug markets in Northern Mexico since 2018. In 2021-2022, study participants described the arrival of new, unusually potent tablets sold as ostensibly controlled substances, without a prescription, directly from pharmacies that cater to US tourists.

**Aims:** To characterize the availability of counterfeit and authentic controlled substances at pharmacies in Northern Mexico available to English-speaking tourists without a prescription.

**Methods:** We employed an iterative, exploratory, mixed methods design. Longitudinal ethnographic data was used to characterize tourist-oriented micro-neighborhoods and guide the selection of n=40 pharmacies in n=4 cities in Northern Mexico. In each pharmacy, samples of “oxycodone”, “Xanax”, and “Adderall” were sought as single pills, during English-language encounters, after which detailed ethnographic accounts were recorded. We employed immunoassay-based testing strips to check each pill for the presence of fentanyls, benzodiazepines, amphetamines, and methamphetamines. We used Fourier-Transform Infrared Spectroscopy to further characterize drug contents.

**Results:** Of n=40 pharmacies, one or more of the requested controlled substance could be obtained with no prescription (as single pills or in bottles) at 28 (70.0%) and as single pills at 19 (47.5%). Counterfeit pills were obtained at 11 pharmacies (27.5%). Of n=45 samples sold as one-off controlled substances, 18 were counterfeit. 7 of 11 (63.6%) samples sold as “Adderall” contained methamphetamine, 8 of 27 (29.6%) samples sold as “Oxycodone” contained fentanyl, and 3 “Oxycodone” samples contained heroin. Pharmacies providing counterfeit drugs were uniformly located in tourist-serving micro-neighborhoods, and generally featured English-language advertisements for erectile dysfunction medications and “painkillers”. Pharmacy employees occasionally expressed concern about overdose risk and provided harm reduction guidance.

**Discussion:** The availability of fentanyl-, heroin-, and methamphetamine-based counterfeit medications in tourist-oriented independent pharmacies in Northern Mexico represents a public health risk, and occurs in the context of 1) the normalization of medical tourism as a response to rising unaffordability of healthcare in the US, 2) plummeting rates of opioid prescription in the US, affecting both chronic pain patients and the availability of legitimate pharmaceuticals on the unregulated market, 3) the rise of fentanyl-based counterfeit opioids as a key driver of the fourth, and deadliest-to-date, wave of the opioid crisis. It was not possible to distinguish counterfeit medications based on appearance of pills or geography of pharmacies, because identically-appearing authentic and counterfeit versions were often sold in close geographic proximity. Nevertheless, drug consumers may be more trusting of controlled substances purchased directly from pharmacies. Due to Mexico’s limited opioid overdose surveillance infrastructure, the current death rate from these substances remains unknown.

## Introduction

Counterfeit pharmaceutical drugs—especially those containing illicitly manufactured fentanyls (IMF)— are playing an increasing important role in the United States (US) overdose crisis^1–3^. IMF and other synthetic opioids have transformed the risk environment for people who use drugs (PWUD) given their much higher potency and shorter half-life compared to other opioids. Although reports of counterfeit prescription opioids containing IMF surfaced as early as the early 2010s,^4^ in recent years they have become commonplace across the US^5,6^. IMF-based pills (e.g., counterfeit “M30s”) have been linked to large increases in overdose mortality on the West Coast especially^7,8^, and have been found in virtually every state in the US^9^. They have also been driving a large relative increase in the overdose death rate of adolescents^3^, who are more likely to experiment with drugs they perceive as prescription pharmaceuticals relative to powders sold as heroin, or other drugs that are more stigmatized^10^.

For the general population, there is a profound added risk of counterfeit prescription medications relative to other fentanyl-based illicit drug formulations. Pills purporting to be pharmaceuticals may be perceived as a lower-risk category of recreational drugs^2^, despite currently representing one of the most potentially lethal options for illicit drug use, especially because they may be more likely to be used by individuals with no tolerance to opioids. Counterfeit versions of psychoactive drugs have been identified in numerous contexts, often using point-of-care techniques like Fourier transform infrared spectroscopy and fentanyl and benzodiazepine testing strips, or more time and cost-intensive techniques like gas chromatography with mass spectroscopy^5,6,9,11–14^. Importantly, the existence of counterfeit versions of (non-psychoactive) medications that are expensive or difficult to obtain is a longstanding and well-described problem in many low- and middle-income countries^15–18^.

Our ethnographic team has conducted longitudinal research focused on illicit drug markets in Northern Mexico since 2018^19–22^. In 2021-2022, study participants—especially those who were US citizens that frequently visited in or stayed in Mexico to consume illicit drugs—began describing new, unusually potent medications, sold ostensibly as controlled substances, from pharmacies that cater to English-speaking tourists. This raised suspicion for the proliferation of counterfeit prescription drugs in brick-and-mortar pharmacies^1^.

Although counterfeit prescriptions have been described in the US illicit drug market—and it is well-known that a large portion of the IMF and methamphetamine that are pressed into counterfeit prescription drugs sold in the US originate in Mexico^23,24^—the prevalence of their consumption among PWUD in Mexico is not well-described in academic literature. Previous drug checking and ethnographic studies have characterized the arrival of IMF to Tijuana and elsewhere in Mexico, especially in the form of China White (ostensibly powder heroin, often found containing fentanyl), which has increased in prevalence in recent years^21,25^. However, previous ethnographic literature describing illicit drug supply chains in Mexico indicate that a complicated and varying set of cartel politics and on-the-ground dynamics limit which products are sold in Mexico versus those exclusively earmarked for export to the US^21,26^. It should therefore not be assumed that illicit drug products produced in Mexico for export to the US—including counterfeit pills—are consumed by PWUD in Mexico.

Given the potential public health risks associated with IMF-based counterfeit prescriptions sold from pharmacies, we sought to characterize the availability and composition of pills sold as ostensibly controlled substances in pharmacies across Northern Mexico.

## Methods

### Ethnographic Methods For Preexisting Data

Ethnographic data were collected as part of a wider study investigating shifting risk environments of PWUD in Mexico. Sampling dynamics and methodology have been described extensively elsewhere^19–22^. Briefly, we targeted initial fieldwork towards the immediate surroundings of drug- and sex-tourism micro-neighborhoods. The largest fraction of participants in the initial ethnographic work were deported individuals who had spent significant time in the US but could not return at-will. However, a sizeable minority of key ethnographic informants consisted of US citizens and residents intentionally visiting or residing in Mexico for the purpose of purchasing and consuming illicit drugs at far cheaper prices than those seen in proximate parts of the US. Ethnographers accompanied and informally interviewed participants as they engaged in routine daily activities, including the acquisition, preparation, and consumption of illicit drugs, and the generation of funds. This anthropological approach allows for the generation of ‘common-sense’ understandings of the risk environment for PWUD, and the generation of novel hypotheses about critical public health risk factors and dynamics. With IRB approvals, we employed a conversational interview format, frequently using audio recording with participant permission. Over time, most key informants were formally and informally interviewed on dozens of occasions during the research process. All ethnographers were bilingual, and textual data were translated to English for presentation. Study protocols were approved by the institutional review board at the University of California, Los Angeles, in the United States, and the institutional review board at Prevencasa, a non-governmental organization, in Baja California, Mexico.

During the process of accompanying key informants, visits to pharmacies were commonplace, especially among US citizens and residents participating in cross-border drug use (visiting for short periods or living in Mexico for the purpose of drug use), who would occasionally purchase one-off benzodiazepine pills, bottles of tramadol for use in heroin cessation, or boxes of syringes at pharmacies proximate to drug and sex tourism microneighborhoods.

### Additional Ethnographic Methods For Current Study

For the present article, longitudinal ethnographic data were used to characterize the tourist-oriented micro-neighborhoods where the study occurred and guide the selection of n=40 pharmacies in n=4 cities in Northern Mexico (of note, we are choosing not to specifically name the cities and to obfuscate any identifying information). Given the existing information available, pharmacies were chosen strategically to be geographically broad—both within and between cities—maximizing the probability of discovering counterfeit prescriptions in at least one area, should they exist. This was accomplished by sampling various pharmacies in distinct types of micro-neighborhoods within several cities (e.g. those focused on drug and sex tourism and those focused on more mainstream tourism.) Therefore the study sought to be hypothesis generating and exploratory, not assessing prevalence in a representative fashion. Based on initial ethnographic insights, all pharmacies appeared to be independent entities, not part of one of the many popular national pharmacy chains. These pharmacies were deemed the most likely to provide counterfeit medications and other controlled substances without a prescription. All pharmacies were located in areas frequented or transited by US-based tourists.

In each pharmacy, samples of controlled substances, initially requested as “oxycodone”, “Xanax”, and “Adderall”, were sought as single pills, during English-language encounters. English-only encounters were used given that preliminary ethnographic results indicated that controlled substances were often sold ‘only for tourists,’ so using Spanish might be associated with a lower probability of identifying counterfeit pharmaceuticals. Generic names (e.g. alprazolam) were used when pharmacy staff did not recognize brand names. Each category of pills was requested as single tablets, and when multiple formulations were offered of the same medication, one sample of each formulation was acquired. Each category of medications was sought in each pharmacy included in the sample, and ‘oxycodone’ was typically the first-medication to be requested in each pharmacy.

Consistent with other studies of ‘real-world’ product availability^27^, pharmacy staff were not informed that a study was occurring. Immediately after each encounter, metadata was recorded, indicating if controlled substances were available in any form (including full bottles), and if single pills could be obtained (which were universally requested when full bottles were offered). Only single pill samples were obtained and analyzed because initial ethnographic insights led researchers to believe that they were widely available, and would represent the highest potential for representing counterfeit products. Labeled bottles of pills were deemed less likely to contain counterfeit substances, and also would have involved considerably increased costs. After each encounter, detailed ethnographic accounts were recorded, transcribed, added to the existing corpus of data, and coded.

### Ethnographic Analysis of Pre-Existing and Newly Collected Data

The final ethnographic database, included previously collected data, as well as new data collected for the current study (as described above) and consisted of more than 100 transcribed recordings, 500+ pages of fieldnotes, 600+ photographs, and dozens of videos documenting practices in natural environments unfolding in real time. Data from prior to 2022 (2019-2021) were drawn from the pre-existing corpus of ethnographic information, whereas novel ethnographic data were collected in 2022 for this study. All qualitative data were entered into NVivo and analyzed for emergent themes. Of particular relevance for this analysis, all encounters occurring in pharmacies were analyzed separately to track the evolving use of pharmacies by PWUD over time (2019 to 2022). Narratives from pharmacy staff were also specifically assessed when spontaneously offered during the course of the study, although they were not actively solicited. They were combined with narratives from key ethnographic informants from the wider study with detailed relevant knowledge regarding medication quality, safety, contents, and origin. After analysis, key thematic elements from the ethnographic results were presented in a narrative style, consistent with our previous work on these topics^19–21,28–30^. In sum, the final set of ethnographic data analyzed consisted of 1) the subset of the previously existing and continuously evolving corpus of ethnographic data (recorded interviews and field notes) from the wider study that pertained to pharmacy-based practices and encounters 2) ethnographic data recorded immediately after the pharmacy-based encounters specifically conducted for this study.

### Drug Checking Methods

All samples were processed in a standardized fashion (see supplement for step-by-step details). Briefly, the entire pill was pulverized using a glass instrument in a single-use plastic receptacle, and pill contents were mixed thoroughly to minimize heterogeneity. A small sub-sample was then selected—the smallest quantity that completely covered the crystal window of the Fourier-Transform Infrared (FTIR) Spectroscopy window. An alpha-2® FTIR spectrometer (Bruker: Billerica, MA) was employed to characterize drug components using a series of libraries describing licit and illicit substances^13^ (see supplement). Subsequently, the same sample was added to 1.0 ml of filtered water, and was agitated manually for ten seconds, followed by agitation using a test tube oscillator for 10 seconds. We then employed 4 immunoassay-based testing strips from BTNX laboratories for each sample, to check each pill for the presence of 1) fentanyls, 2) benzodiazepines, 3) amphetamines, and 4) methamphetamines. Each strip was inserted into the dissolved solution for ten seconds, and the result was read after 5 minutes by two trained investigators. In the rare case of disagreement between investigators, or an inconclusive or invalid result, a second strip was employed, providing a definitive result in all cases. For samples sold as Adderall, we further diluted the solution using 120 mL of water for fentanyl testing, to avoid the known issue of false positives at high concentrations of certain stimulants^31^. See the supplement for more details of the drug checking analysis, including the logic employed to reach each final read. Pill samples were determined to be ‘counterfeit’ when it could be demonstrated by immunoassay testing strip or FTIR spectroscopy that they contained a psychoactive ingredient that was not advertised (e.g. containing fentanyl when sold as oxycodone, or containing methamphetamine when sold as ‘Adderall’). Pills were determined to be ‘presumed authentic’ when no inconsistency between pill contents and sold-as status could be determined. Importantly this would not preclude that ‘presumed authentic’ medications could be counterfeits with the correct active ingredients at incorrect dosages, or contain active ingredients that fell outside of the detection capacities of the methods employed here.

## Results

### Drug Checking Results

Of n=40 pharmacies assessed in 2022, the requested controlled substances could be obtained in any form with no prescription, during English-language encounters, at 28 pharmacies (70.0)%, and as single pills at 19 (47.5%) (Figure 1). In n=9 pharmacies, single tablets were not available, but bottles of at least one controlled substance could be purchased without a prescription, including generic alprazolam (US trade brand name Xanax), oxycodone, stimulants, appetite suppressants, and other medications. Counterfeit pills were obtained at n=11 (27.5%) of pharmacies, including methamphetamine-based pills sold as “Adderall” at n=7 (17.5%) of pharmacies and fentanyl-based pills sold as ‘oxycodone’ products at n=6 (15.0%) of pharmacies.

**Figure 1.**
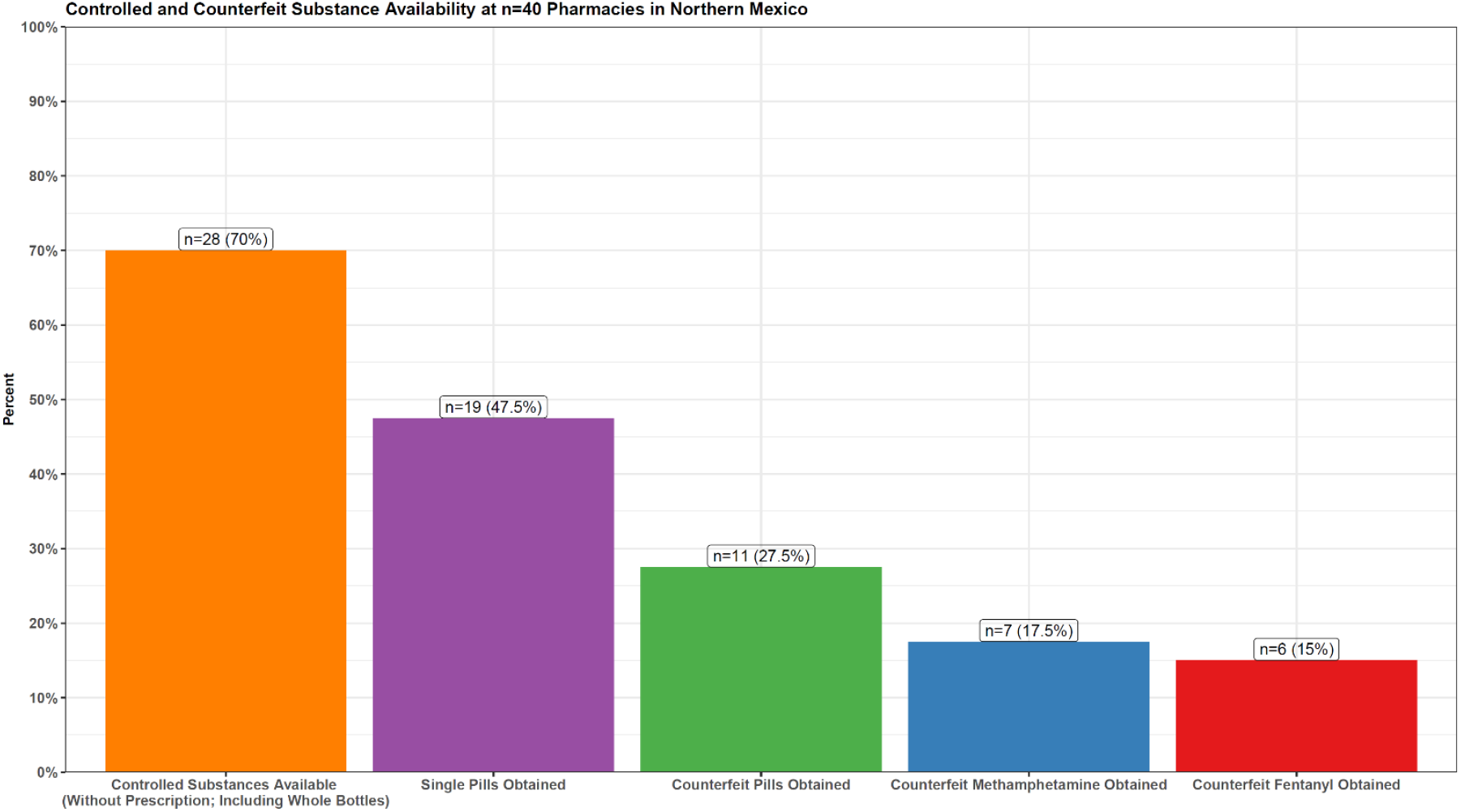
Controlled and Counterfeit Substance Availability at n=40 Pharmacies in Northern Mexico. Pharmacy-level statistics are shown, depicting the availability of controlled substances, as well as counterfeit status as determined with immunoassay and FTIR spectroscopy. Data from n=40 pharmacies are included.

Of n=45 samples sold as the requested controlled substances, 27 were oxycodone, 11 were Adderall and 7 were Xanax. N=18 (40.0%) were determined to be counterfeit including 7 of 11 (63.6%) samples sold as “Adderall” that contained methamphetamine, 8 of 27 (29.6%) samples sold as “oxycodone” that contained fentanyl, and n=3 (11.1%) “oxycodone” samples that contained heroin (Figure 2). None of the pills sold as Xanax were found to be counterfeit. One sample sold as “Vicodin” was shown to contain only lactose and tramadol on FTIR spectroscopy (note: authentic Vicodin contains hydrocodone, not oxycodone. However, when pharmacy employees offered other types of prescription opioids in response to “oxycodone,” they were acquired and processed, given that many tourists may not recognize the difference). A wide variety of presumptively authentic controlled substances (based on fentanyl and methamphetamine negative status with immunoassay strips, and FTIR confirmation) were available (see Figure 3). A variety of phenotypes of counterfeit medications were also observed (Figure 3). See the supplemental materials for more details regarding how final drug checking designations were determined.

**Figure 2.**
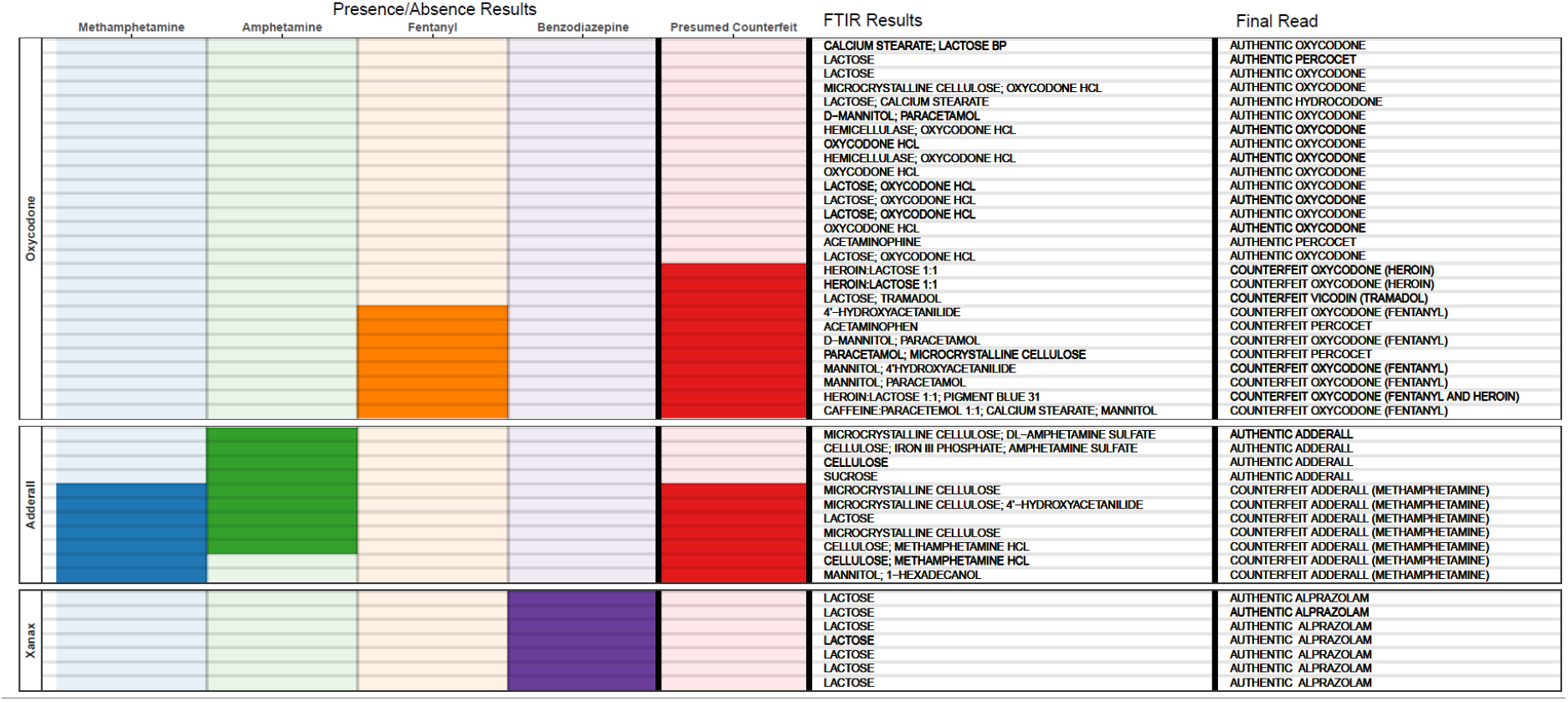
Drug Checking Results From n=45 Pill Samples Obtained in Northern Mexico. Pill-level data are shown, with one row per sample analyzed. The labs on the far left show the prompt used to obtain each pill, i.e. what the sample was sold as. Binary, presence/absence results are shown for four drug categories using shaded vs unshaded boxes. FTIR results are shown in free text, with up to 3 results separated by semi-colons. The ‘final read’ as determined by the investigators is shown in free text. Not all final reads are 100% definitive, rather it represents the most likely result as determined by all forms of drug checking and ethnographic data (see the supplement for a more elaborate description).

**Figure 3.**
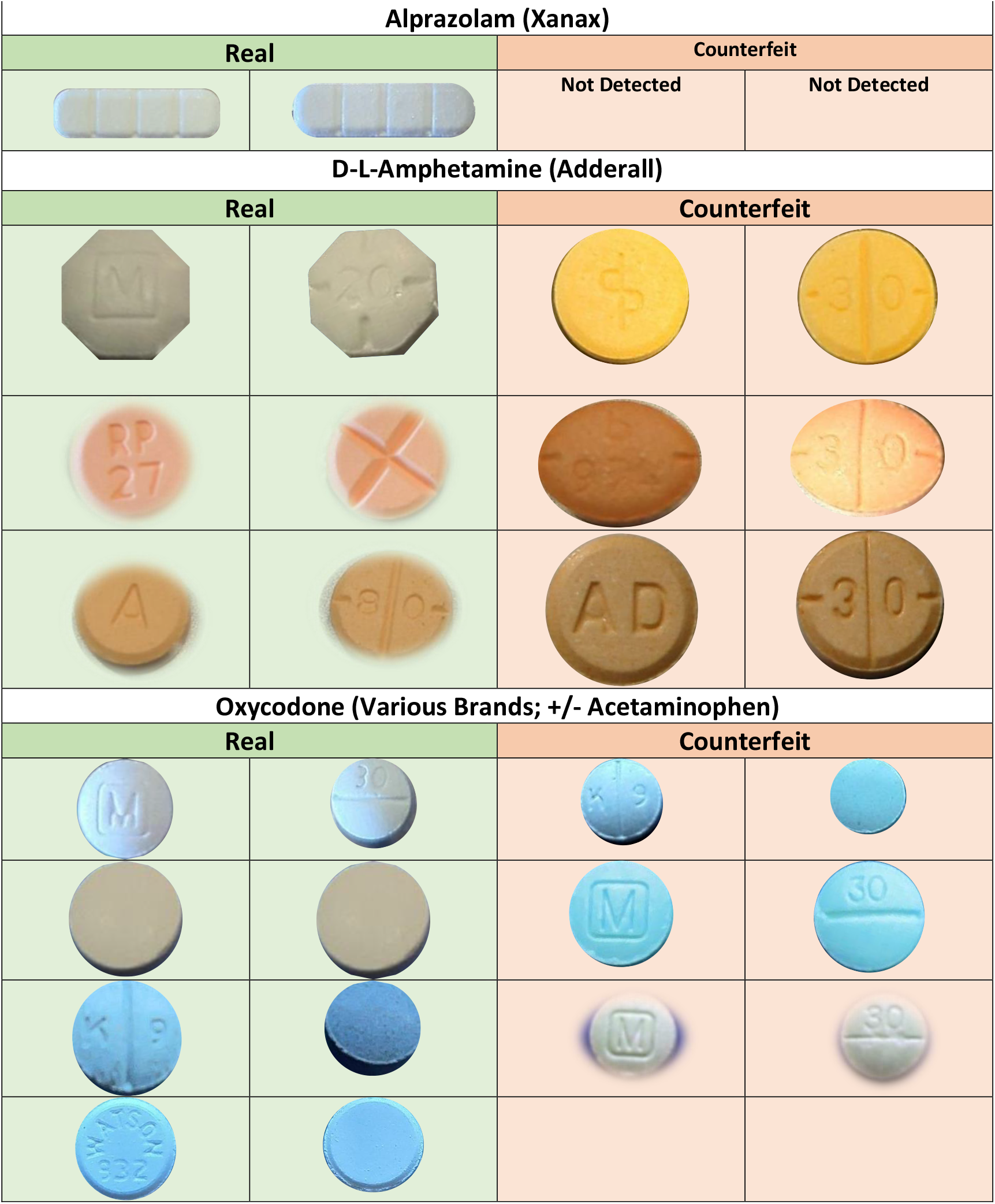
Examples of Known Counterfeit and Presumed Authentic Samples. Photos (front and back) are shown of example pills, by what the sample was sold as, as well as presumed authentic or counterfeit status.

Despite intensive fieldwork employed in concert with drug checking technologies, the ethnographic team determined that at the independent pharmacies assessed here, it was not possible to distinguish counterfeit medications from their authentic counterparts based on appearance, as identically-appearing authentic and counterfeit versions were often sold in close geographic proximity. For instance, pills appearing to be blue “M30” oxycodone tablets were found in authentic, heroin-, and fentanyl-based formulations. Geographic context was at times helpful—with substances sold in specific microneighborhoods found to be more likely to be counterfeit—but provided no guarantee of authenticity. Only the use of several concurrent drug checking technologies provide a reasonable measure of confidence in medication composition. Of note, heroin-based counterfeits were not initially detected by immunoassay strip testing and were only identified by FTIR spectroscopy—a level of drug checking sophistication currently unavailable in many settings where illicit drugs are purchased and consumed.

### Ethnographic Results

*[Fieldnote excerpt from 2019] I’m spending the day walking around the city with Linda (pseudonym used here and throughout the text) a gringa who’s been living in Mexico for a few years after being trafficked, escaping her captors, and who now works mainly as a self-employed sex worker. In her early 30s, she has been dependent on injection opioids for nearly a decade. She uses about 5, 50-peso [$2*.*5 USD] bags of China White heroin (which she believes contain fentanyl) per day and dabbles with many other substances. She is excited to show me a dizzying array of drug consumption spaces in the city. One of the stops is a somewhat formal looking brick-and-mortar pharmacy. “There’s this crazy pharmacy right over here, where you can go in there and buy heroin and cocaine and the fucking white-coat pharmacist sells whatever it is to you! And then you can even take it in the back and use it back there. I have $3, that’s enough, I can show you”. We go into this pharmacy, which I had never taken a second look at, despite passing by it frequently. She greets the pharmacist, who is wearing a white coat, and is visibly covered in tattoos on all exposed parts of his hands, arms, and neck. They speak comfortably in English, and it’s clear they know each other. He barely notices me, which I imagine is because Linda is a charismatic force of nature, who frequently can be seen pulling male clients around in her orbit as she traverses the urban landscape. She hands over 50 pesos and asks for a Valium. He gives her a single pill and 20 pesos in change, and we go into the back room so she can snort it. The room is basically a large closet, and it has a bunch of ride share scooters that look like they’ve been stolen from the US (because they are from companies that don’t operate in Mexico), as well as some bicycles. After she crushes up the pill with a plastic card, and snorts the white powder, she tells me that with the remaining twenty pesos she can show me a shooting gallery right across the street; if we can find five more pesos we can get a 25 peso bag of meth. We leave the pharmacy, cross the street, and duck into an alleyway*…

The ethnographic passage above—from the pre-existing ethnographic corpus—details a pharmacy-based drug acquisition encounter occurring in 2019. Single tablets of controlled substances—especially benzodiazepines—could be routinely obtained at affordable prices from specific pharmacies known to PWUD. Most were proximate to drug- and sex-tourism microneighborhoods catering to English-speaking tourists and heroin- and methamphetamine-dependent Mexican nationals. On rare occasions, the ethnographic team also observed methamphetamine and heroin purchased directly from white coat-clad pharmacy employees. More central to the lives of most PWUD was the acquisition of individual sterile syringes (a legal practice); PWUD often shared details with one another from a complicated taxonomy of which pharmacies would sell syringes to tourists, which ones were open to individuals who appear to have a homeless habitus, and if a cover story was required e.g. “I need syringes for my mom’s insulin”. Oxycodone was not routinely pursued at pharmacies by most ethnographic study participants, and most opioid users used 50-peso (2.5 USD) bags of powder heroin (known as China White) as their main opioid product. On several occasions, study participants bought large bottles of over-the-counter tramadol capsules to facilitate “kicking” their heroin habit.

In 2021-2022, ethnographic participants began describing new, unusually potent controlled substance tablets sold from pharmacies that cater to English-speaking tourists (mostly from the United States):

*[interview recorded in 2022]”I’ve been buying oxy for years here, it used to be the OG [original] oxys only. They were pretty expensive, so I’d only buy em once in a while. But about a year ago, all of the sudden, it was just like ‘boom’ and we had these really strong Oxys for only 20 dollars per pill. So I started doing a bunch of em, like 7 a day if I had the money. But they felt different, the oxys felt like heroin to me, but these new ones, are like fentanyl or some shit. It’s different. They’re stronger but it’s not the same. It used to be more than a dollar per milligram, like 35 dollars for an M30, but all the sudden they were 20 dollars. And then all the pharmacies in this area were selling them. But it’s only for tourists. They’re not supposed to be selling them to locals*.*”*

The ethnographic team observed that in pharmacies offering single tablets of controlled substances, it was commonplace for several distinct presentations of ‘oxycodone’ to be available. In these instances, a broad variety of taxonomies were used to describe the various options available. A very common taxonomy involved describing one set of products as “American” and the other as “Mexican”:

*[field note from 2022] We head into the pharmacy and ask for Oxy. The pharmacy employee flashes us a smile and says “I have Mexican Oxy or I have American Oxy. American Oxy is 35$ for 20mg, and Mexican Oxy is 20$ for 30mg*.*” “Why is the Mexican Oxy stronger and cheaper?” I ask. “Oh the Mexican oxy is very strong, but it’s cheaper because they give it to us for cheaper” he says. “You should only take half, and even that’s going to be a lot. The full one might be too dangerous*.*” I say, “Okay, we’ll take the Mexican Oxy”. He goes under the counter and pulls out a cardboard box full of syringes. He reaches underneath the needles, and pulls up this false bottom on the box, and the bottom is full of these little blue pills, just loose in the box. He takes one out of the pile and puts it in a little plastic bag for us. As he hands it to me. He’s says, “okay guys, these are really strong! Please be careful”. Then we ask about Mexican Adderall. He says “This stuff is cheap, so you have to buy the whole pack”. He shows us the bottle and it says methylphenidate (Ritalin). “So it’s not Adderall” I ask, He says “no, but it’s similar*.*” I ask to see the “American Oxy” and he pulls out a blister pack of circular, white, unmarked tablets. The back of the package reads ‘oxicodona’*.

In this instance, so-called ‘Mexican Oxy’ was fentanyl-positive and determined to be an IMF-based counterfeit product by the drug checking team, whereas ‘American Oxy’ (idiosyncratically labelled in Spanish, and almost certainly of Mexican origin) was determined to be presumptively authentic, confirmed to contain oxycodone on FTIR spectroscopy. Nevertheless, on other occasions, various products sold as “American” and “Mexican” were observed to be both counterfeit and authentic, and no reliable pattern could be discerned. On rare instances, pharmacy employees were more forthcoming about what they suspected were the contents of their medications.

*[field note from 2022] I went into the pharmacy and two young women were sitting behind the counter. I asked for Oxy, and one of them pulled out this plastic case from beneath the counter, with lots of little boxes, like for fishing tackle. It was transparent, so we could see all these different looking pills in little bags. They had two different colors of pills that looked like oxy M30s, one blue and one green. They also had a bunch of white and yellow pills of various sizes and shapes, that looked like Percocet, Norco, etc. I asked, ‘what’s the difference between the green and blue m30s’? One spoke better English and relayed the question to the other in Spanish. They discussed it for a while between themselves, and then after a few moments told us ‘the green one is more like…*.*fentanyl, and the blue one is oxycodone’*.

In this instance, the green M30 tablet sold for $20 USD was confirmed as an IMF-based counterfeit product, whereas the otherwise identical-looking blue M30 tablet sold for $35 USD was found to contain heroin based on FTIR spectroscopy data. A tablet determined to be authentic oxycodone/acetaminophen (sold as “Percocet”) was also obtained during the same encounter.

Pharmacies providing counterfeit drugs were uniformly located in overtly tourist-serving microneighborhoods, and generally featured English-language advertisements for erectile dysfunction medications and ‘painkillers.’ At times pharmacy employees at pharmacies in these areas would state that controlled substances were not sold at their business. These pharmacies tended to be larger, with more employees working at a given time, and located in areas catering to more formal kinds of tourism (i.e. not focused on sex and drug tourism). However, it was not possible for investigators to reliably predict with certainty which pharmacies would sell controlled substances or counterfeit products. On numerous occasions, two pharmacies directly adjacent to one another would provide highly discordant products. Further complicating these dynamics, some pharmacies sold a mixture of counterfeit and authentic oxycodone products.

One-off counterfeit and authentic controlled substance tablets in pharmacies were observed to be stored and accessed in a variety of fashions. However, the degree of discretion employed by pharmacy staff did not appear to be predictive of product authenticity. Tablets were typically stored inside of small plastic bags, kept inside metal breath mint containers in fanny packs, in small cardboard boxes previously containing electronics, in plastic organizer boxes with numerous compartments, or occasionally loose in drawers. At times these boxes were transparent and were left out on pharmacy counters for extended periods of time, as pharmacy staff attended to other patrons seeking medications that were not controlled substances. On other instances, pharmacy staff appeared distinctly concerned about security and employed maneuvers to minimize risk:

*[field note from 2022] I asked for ‘painkillers’, he didn’t understand that word. “like oxycodone” I said, “do you have oxy?” He paused, and looked at me, and then finally said, “yes, we have Mexican oxy” it was 30 USD per pill, they also had Adderall for 25 USD per pill. When I asked about Xanax, he was only going to sell by the bottle, minimum 30 pills. He did the math and took the money for everything I had asked for. I didn’t see that he had contacted anyone, but somehow, he must have relayed my order. But after he took the money, he just kind of stood behind the counter, counted the money and just like leaned on the wall, like nothing was going to happen. So I thought, “are we about to get ripped off?” Then I noticed there were a few other people in the store, there were a few other employees, they were watching us pretty closely, and it felt like they were blocking the door. In hindsight, that was probably because there were quite a few police cars right outside in that area. It was at least two full minutes of just standing around, and then someone from the outside finally came in, and the guy moved around me to the back of the store in this really awkward way and pulled the little baggie with two pills out of his pocket and handed it to me and I left. It felt like way more precautions were taken than other encounters, which I attributed to it being in a part of town with more formal tourism in the area, so they took more precautions than pharmacies in the part of town where people go for sex tourism, where things were a bit more out in the open*.

Pharmacy employees selling counterfeit tablets occasionally expressed concern about potency and provided harm reduction guidance, classically “only take half and see how you feel” (see third ethnographic passage above). Additionally, on several occasions, pharmacy employees selling exclusively authentic oxycodone products would counsel caution purchasing products elsewhere, implying risk of overdose or adverse drug reactions:

*[field note from 2022] We head into this small pharmacy which caught our eye because it says “English Spoken” in big letters on the front, which at this point was starting to seem like code for “we have recreational drugs”. The woman behind the counter didn’t speak Native English, but she spoke pretty good casual English. We ask “do you have Oxycodone?” and she says “Yes”, very matter of fact, and then starts cracking jokes about how much better her supply is than the neighboring pharmacies. She says “look guys, if you get oxys in most of the places around here, they’re not real, it’s just going to be fentanyl. And look, we want you to have a good time, but if you take one of those, you are not coming back. OK? You’re not coming back and we don’t want that. But I have the real thing. See I’m going to show you the package. Always ask to see the package, OK?”. She pulls out a blister pack that is half-full with about 10 white circular pills, which reads “oxicodona, 20mg” on the back. “These are the only real oxys around, OK? I only have 20 milligrams. I used to have 40s and 80s and they’re impossible to get now. But you guys are gonna be happy. I guarantee you’re gonna come back!” she exclaimed laughing*.

In this encounter, the 20mg oxycodone pills were determined to be presumed authentic. Key informants also confirmed that consumption of counterfeit IMF-based tablets, often pressed to look like blue Oxycodone M30s, was subjectively associated with an increased risk of overdose:

*[Interview from 2022] Interviewer: “Have people been overdosing here on the M30s?” Participant: “Yeah, well I’ve never seen a tourist die from one, but they always do them in the hotel, so I’m not sure. But one of my homies did almost, yeah, he was smoking the blues and he nodded real hard, actually right over here, and the girl from the pharmacy had to keep pushing on his chest like this (motions doing chest compressions)*.

*Interviewer: “Did they call 9-1-1?”*

*Participant: “Yeah, they came, and they saved him, but it was close. I had never seen someone get that close from the OG oxys”*

A distinct phenotype of tourist-oriented pharmacies—focused on selling controlled substances exclusively in bottles and blister packs of quantities ranging from 10 to 150 tablets—was noted in several locations. Pharmacy employees in these locations often had lists or ‘menus’ of drug options— sitting in plain sight or posted on the outside of the store—(spanning from benzodiazepines, muscle relaxers, amphetamine-based diet pills, and anabolic steroids, among others), as well as cellphone photos of bottles that were displayed to clients upon request (see Figure 4). Each bottle of pills sold in these locations was typically priced at $200-$400 USD. Pharmacy employees offered various strategies for successful importation of the medications to the US on return flights, and some offered to facilitate international shipping for an additional fee. At a subset of these pharmacies, single pills could be obtained, but only after considerable insistence that a large quantity was not of interest.

## Discussion

Leveraging recent improvements in point-of-use drug checking technologies, we provide the first characterization—to our knowledge—of the contents of medications sold at pharmacies in tourist-serving areas of Northern Mexico, in single pill form, to English-speaking tourists without a prescription. We find a high rate of counterfeit products, with widespread fentanyl and methamphetamine prevalence in numerous sites.

The availability of fentanyl, methamphetamine, and heroin-based counterfeit medications in Northern Mexican pharmacies that are oriented towards serving tourists represents a distinct public health threat. These medications have been implicated in large increases in overdose risk in the United States, especially among subpopulations of individuals that are willing to experiment with prescription pills but not more stigmatized formulations like powder heroin^2–6^. Although IMF-based pills represent a very high-risk category of illicit drug product, drug consumers may be more trusting of controlled substances purchased directly from pharmacies. Critically, it is not possible to distinguish counterfeit medications based on appearance of pills or geography oh pharmacies, as identically-appearing authentic and counterfeit versions are often sold in close geographic proximity. Harm reduction logic would dictate that a person consuming purported controlled substances purchased at pharmacies in these micro-neighborhoods should test each pill on each occasion that drugs are consumed, to ensure IMF and methamphetamine contamination has not occurred.

The presence of controlled substances in pharmacies of northern Mexican occurs in the context of a long history of drug and medical tourism to Mexico by US residents and citizens^32–36^. There is a well-established practice of US tourists traveling to Mexico to purchase medications at a far lower cost than those available in the US, and often with no prescription, for medications that require a formal prescription and potentially expensive doctor’s visit in the US. This demand in large part reflects the extremely expensive, unaffordable, confusing, and exploitative nature of the US healthcare system, where many individuals fear that even simple healthcare encounters may result in financially catastrophic outcomes^37^. Additionally, it is well-described in the literature that many prescription drugs are dozens to hundreds of times more expensive in the US than in other countries, including Mexico^38^. Legally, this does not apply to controlled substances—such as opioids, benzodiazepines, or stimulants— which Mexican law dictates do require a special kind of prescription from a licensed physician authorized to prescribe psychoactive drugs^39–41^. However, in specific locations we observed a widespread practice wherein certain kinds of controlled substances, especially alprazolam, were readily available with no prescription, at pharmacies that visibly cater to English-speaking tourists. Indeed, it was never the case in any of the tourist-oriented pharmacies where controlled substances were obtained that a prescription was required; specific medications were either available or unavailable, regardless of prescription status.

In some of these pharmacies, employees routinely offered advice to English-speaking tourists on how they can smuggle controlled substances back into the United States and avoid detection. In this context, it could be especially difficult to recognize the threat of possibly counterfeit controlled substances— because a mix of counterfeit and authentic controlled substances are illegally sold (either by the lack of a legally-required prescription, or by being illicit drugs) from the same locales. For an English-speaking tourist with a poor level of knowledge of the Mexican legal landscape, it may not be immediately apparent that the sale of any controlled substance without a special prescription constitutes an illegal act. The sale of individual pills from larger bottles or boxes is also not legally permissible.

Additionally, it is important to note that the rate of opioid prescription in the US has fallen drastically over the past decade, by nearly 50% in terms of morphine milligram equivalent (MME), between 2010 and 2020^42^. These decreases have been shown to have affected many patients with known painful chronic conditions, including terminal cancer, and other palliative care patients^43^. Many patients have been rapidly tapered off opioid regimens, which has been associated with increased rates of suicide and drug overdose^44,45^. We would argue that a large unmet demand for diverted and legitimate prescription opioids has likely led to widespread consumption of counterfeit opioids in the US by witting and unwitting consumers. Similarly, recent shortages of Adderall have led to substantial unmet demand for amphetamine among US patients and diverted medication consumers, which some drug policy experts have hypothesized may lead to increased use of counterfeit methamphetamine-based Adderall tablets^46^.

The rise of counterfeit pills in the US, as well as the shifts we note here, seem to have intensified during the COVID-19 pandemic, which may have driven these trends in unknown ways. Disruptions to the illicit drug supply during the pandemic may be involved, and this represents an important area for further study.

Due to Mexico’s limited opioid overdose surveillance infrastructure, the current death rate from these substances remains unknown. There is a lack of drug mortality surveillance data in Mexico, largely stemming from limitations on epidemiological and drug checking data sources. Toxicological and autopsy data are extremely limited^47^, and most overdose deaths are coded with non-specific codes such as ‘cardiac arrest’ which do not indicate the true underlying cause of mortality^47,48^. A number of qualitative and drug checking studies have indicated that fentanyl has arrived to Tijuana and other northern Mexican border cities^21,22,25^. Yet the quantitative epidemiological impact of these shifts, and any further implications from the availability of counterfeit medications, has not been adequately characterized.

### Limitations

This study is exploratory in nature, and the results should be considered hypothesis-generating and limited, requiring validation. Importantly, we did not seek to characterize or represent the prevalence of counterfeit medications across all pharmacies in the four cities of interest. Instead, we used ethnographic data to guide a purposive sampling approach that we believed was most likely to document the presence of counterfeit medications if they were present in any one of an array of intentionally selected micro-neighborhoods. This study also leverages several drug checking methodologies that are relatively new, and which require validation. Although we took extensive efforts to reduce false positives and negatives (see supplement), we cannot rule them out, and all results should be interpreted in light of the inherent uncertainties of modern drug checking methodologies. We also were not able to employ gas chromatography, mass spectrometry confirmatory testing for the samples analyzed here, although that has been done in some similar analyses. We also did not seek to characterize the full population of individuals that may be purchasing these drugs, relying on a convenience sample of known informants. Data came from a single region of Mexico, and from independent, not corporate pharmacies, and therefore do not represent the full market of pharmacies.

## Conclusions

The availability of fentanyl-, heroin- and methamphetamine-based counterfeit medications in tourist-oriented independent pharmacies in Northern Mexico represents a public health risk to Mexican residents and tourists, and occurs in the context of 1) the normalization of medical tourism as a response to rising unaffordability of healthcare in the US, 2) plummeting rates of opioid prescription in the US, affecting both legitimate pain patients and the availability of legitimate pharmaceuticals on the black market, 3) the rise of fentanyl-based counterfeit opioids as a key driver of the fourth, and deadliest-to-date, wave of the opioid crisis. Among the samples obtained from the tourist-oriented independent pharmacies we studied here, it was not possible to distinguish counterfeit medications based on appearance of pills, because authentic and counterfeit versions are often sold in close geographic proximity and are visually and otherwise indistinguishable from one another. Nevertheless, English-speaking tourists may be more trusting of controlled substances purchased directly from pharmacies. Due to Mexico’s limited opioid overdose surveillance infrastructure, the current death rate from these substances remains unknown.

## Data Availability

All data used in the paper are contained in the manuscript's supplement.

## Acknowledgements

We gratefully acknowledge the contributions of Dr. Pamina Gorbach and Dr. Steven Shoptaw to earlier drafts of this manuscript, as well as those of several collaborators at Mexican institutions who chose to remain anonymous. The authors wish to thank the UCLA Center for AIDS Research under U.S. National Institutes of Health (grant P30AI152501) and the UCLA AIDS Institute for use of the FTIR spectrometer, made possible through generous support from the James B. Pendleton Charitable Trust. CLS was supported by the U.S. National Institute on Drug Abuse (grant K01DA050771). AB was supported by the U.S. National Institute on Drug Abuse (grant NIDA DP2 DA049295). DGM was supported by the U.S. National Institute on Drug Abuse (grant K08DA048163). JRF was supported by the National Institute of General Medical Sciences (grant GM008042). The funders had no role in study design, data collection and analysis, decision to publish, or preparation of the manuscript.

