## Supplement for "Fentanyl, Heroin, and Methamphetamine-Based Counterfeit Pills Sold at Tourist-Oriented Pharmacies in Mexico: An Ethnographic and Drug Checking Study"

**Steps Taken To Conduct The Study**

The following steps detail the process by which samples were chemically analyzed using a Bruker Alpha II FTIR spectrometer equipped with the platinum attenuated total reflectance (ATR) module, which has an ATR diamond crystal. This machine was operated using the OPUS spectrometer software, a program used to match an unknown sample’s absorbance spectrum with reference library spectra.

1. Samples were stored in individual plastic receptacles to prevent cross contamination.
2. At the beginning of each testing clinic the FTIR spectrometer and a dedicated laptop were placed on a sturdy table in a well-ventilated area.
3. The FTIR spectrometer was set-up by connecting the basic module, which houses the interferometer, beamsplitter, IR source, and other optical components, to the sampling module. The lock on the basic module is pressed down and the sampling module is slid into place. Finally, the basic module lock is pressed again, locking the sampling module in place.
4. An Ethernet cable was used to connect the spectrometer to the laptop. Both the laptop and spectrometer were plugged into an external power supply.
5. After turning on the PC, The OPUS DrugID Wizard interface was opened, automatically starting the initial warm-up calibration. Following this, the spectrometer will run a performance test.
6. Since our spectrometer moves between different clinic sites, we also ran a Performance Qualification (PQ) test after the initial calibration process. If the spectrometer stays in a temperature-controlled benchtop location and is not unplugged this would instead be done once a week. The PQ test was initiated by double-clicking on the green circle in the bottom right-hand corner of the OPUS interface, prompting the Instrument Status box to appear. The technician then double clicked on the ATR-Diamond box with the FTIR spectrometer image. The technician then selected the PQ checkbox and pressed the Run Tests button. A small dialog box then appeared stating the instrument test is running with the bottom task bar showing a green progress bar. After the PQ test finished, a PDF entitled OVP-PQ Test Protocol appeared, detailing the test result. If the test failed, the PQ test was run again.
7. Once the spectrometer was ready to use, each technician was equipped with a KN95 mask and nitrile gloves to handle samples.
8. Using a glass instrument and a single-use plastic receptacle, the entire pill was pulverized. Pill contents were mixed thoroughly to minimize heterogeneity.
9. Between each sample preparation and subsequent chemical analysis, the metal tray was thoroughly cleaned twice with isopropyl alcohol and Kimwipes and technicians changed their gloves to prevent cross contamination.
10. Before chemical analysis, a background spectrum was measured to reduce the effects the FTIR spectrometer itself has on the sample’s spectrum.
11. A disposable plastic instrument was used to place the minimum amount of sample to cover the window on the sampling module, about 5mg.
12. A small square of aluminum foil was placed on top of the sample powder to protect the diamond crystal and the diamond crystal was gently clamped on top.

13. The Measurement command was selected, prompting a dialog box to open.
14. The Start Sample Measurement button in the dialog was clicked.
15. After the sample's spectrum was generated, the technician then clicked the Evaluate tab and selected the Spectrum Search option.
16. Once the Spectrum Search pop-up appeared, the Search Parameters were selected. Our team used the Vector Normalization method and First Derivative for the spectrum correlation. The minimum hit quality listed was 100 and the maximum number of hits, or matching reference spectra, was set at 30.
17. Then the technician clicked the Select Libraries tab on the Spectrum Search pop-up, choosing four reference libraries that contain spectra from commonly seen drugs and adulterants: TICTAC ATR-FTIR Drug Library, the British Columbia Centre on Substance Use (BCCSU) ATR-FTIR Library, ATR-LIB-PHARMA-1 Library, and ATR-LIB-PHARMA-2 Library. If a substance was not listed in a selected reference library, it would not be able to be detected, which is why we utilized four robust libraries.
18. The technician then selected the Search Library button at the bottom of the pop-up window and compared the sample spectrum, which is a sum of all the spectra of the sample components, to the suggested reference spectra.
19. After a component was identified, it was autosubtracted from the sample spectrum by right clicking on the component name in the list of suggestions to identify remaining components.
20. Spectral analysis of samples is difficult beyond four components, so we recorded up to three results.

The limit of detection for the FTIR spectrometer is about 5% by mass. Since some drugs may produce significant effects below this level, and the FTIR can only confirm the presence of compounds and not their absence, we employed other drug checking techniques to further characterize the samples. To identify the presence of fentanyls, benzodiazepines, amphetamines, and methamphetamine, we used BTNX Rapid Response lateral flow immunoassay test strips, which were originally intended for the detection of illicit drugs in urine samples. These strips may detect the presence of a drug but cannot identify the drug in each class that crossreacted with the antibody. The sample is unaltered during analysis with the FTIR machine, so we test the same sample that the spectrometer analyzed in these assays.

**Test Strip Procedure**

1. 1 mg of the same sample and 1.0 ml of filtered water were added to an Eppendorf vial. The vial was agitated manually for ten seconds followed by agitation using a vortex mixer for 10 seconds.
2. We then employed 4 immunoassay-based testing strips from BTNX laboratories for each sample, to check each pill for the presence of 1) fentanyls, 2) benzodiazepines, 3) amphetamines, and 4) methamphetamine. The wavy side of each strip was inserted into the dissolved solution for ten seconds, taking care to not insert it past the thick blue max line, and placed flat on a clean, non-absorbent surface to allow the solution to run up the strip via capillary action.
3. After 5 minutes, two trained investigators read the test strip results. In the rare case of disagreement between investigators, or an inconclusive or invalid result, a second strip was employed, providing a definitive result in all cases. For samples sold as Adderall we further

diluted the solution using 120 mL of water for fentanyl testing, to avoid the known issue of false positives at high concentrations of certain stimulants.[3]

4. Interpretation of test strip results: One line is positive, two lines are negative, and the test is invalid if no lines show or there is no control line. If a fentanyl analogue was identified on the FTIR spectrometer, a fentanyl test strip was not run.

**Reagents Procedure**

To differentiate between amphetamine and methamphetamine, Simon's reagents were utilized on samples where both amphetamine and methamphetamine immunoassay test strips were positive. Simon's reagents consist of solution A, a mixture of 2% sodium nitroprusside and 2% acetaldehyde and solution B, a mixture of 2% sodium carbonate.

1. Wearing nitrile gloves and a mask, a technician placed a grain of sample onto a ceramic plate.
2. The technician then added a drop of solution A followed by a drop of solution B onto the plate.
3. If a secondary amine group was present, as is the case in methamphetamine, the technician would note the color change to a blue solution.

**Sample Disposal Procedure**

To dispose of the samples, we used nontoxic DisposeRx powder to render the drug samples inert in a viscous gel.

1. The sample powder, a DisposeRx packet, and 15 mL of filtered water were added to a 22 mL vial.
2. The vial was then agitated manually until the gel formed.
3. This vial was then safely discarded into the trash.

**Drug Checking Considerations (Useful in Understanding 'Final Read')**

***BTNX Fentanyl Test Strips***

The cut-off concentration in which the fentanyl test strips detect fentanyl and select analogues in a sample is 200 ng/mL (i.e. they are extremely sensitive).[1,2] BTNX fentanyl test strips cross react with Acetyl Fentanyl, Butyryl Fentanyl, Carfentanil, Fentanyl, p-Fluoro fentanyl, Furanyl Fentanyl, 3-Methyl Fentanyl, Ocfentanil, Remifentanil, Sufentanil, and Valeryl Fentanyl.[1,2]

False positive results on BTNX fentanyl test strips that indicate the presence of fentanyl are a known occurrence with methamphetamine, MDMA, and diphenhydramine at concentrations at or above 1 mg/1mL due to the cross reactivity of the antibody for these other substances.[3,4] When these drugs and adulterants may be in high concentrations, such as in the drugs sold as Adderall, our protocol was to add 120 mL of water to the sample solution to prevent this cross reactivity and a subsequent false positive which is a successfully used technique at other drug checking clinics.

***BTNX Benzodiazepine Test Strips***

Benzodiazepine test strips have a higher limit of detection than fentanyl test strips, so more of a sample is needed to reliably detect benzodiazepines. Each benzodiazepine also has a different concentration in which it causes a positive result with a cut-off concentration of 300 ng/mL.[5] False negatives are more prevalent due to these considerations.

| **Benzodiazepine Test Strip Cross Reactivity List[5]** | |
|---|---|
| **Benzodiazepine** | **Concentration in which it causes a positive result on immunoassay strip** |
| Oxazepam | 300 ng/mL |
| Alprazolam | 125 ng/mL |
| Bromazepam | 625 ng/mL |
| Chlordiazepoxide | 2500 ng/mL |
| Clobazam | 63 ng/mL |
| Clonazepam | 2500 ng/mL |
| Clorazepate | 3300 ng/mL |
| Desalkflurazepam | 250 ng/mL |
| Diazepam | 250 ng/mL |
| Estazolam | 5000 ng/mL |
| Flunitrazepam | 375 ng/mL |
| Flurazepam | >100,000 ng/mL |
| Lorazepam | 1250 ng/mL |
| Lormetazepam | 1250 ng/mL |
| Medazepam | >100,000 ng/mL |
| Midazolam | >100,000 ng/mL |
| Nitrazepam | 25,000 ng/mL |
| Norchlordiazepoxide | 250 ng/mL |
| Nordiazepam | 500 ng/mL |
| Prazepam | >100,000 ng/mL |
| Temazepam | 63 ng/mL |
| Triazolam | 5000 ng/mL |

**Simon's Reagents**

Simon's reagents were selected for further chemical analysis to distinguish between amphetamine and methamphetamine by visualizing a color change. Amphetamine and methamphetamine are structurally similar, with the addition of a methyl group attached to the amine group in methamphetamine. The Simon's reagents react with this secondary amine, but not the primary amine of amphetamine, turning the sample blue.[6] Other drugs such as MDMA may also contain secondary amine groups that would react with Simon's reagents, so this step was completed after the FTIR and immunoassay strip analysis steps to prevent false positive results.

**Pharmaceutical Adderall Test Results**

As a sensitivity analysis, and to inform the main study findings, based on client serving drug checking services, samples of Adderall prescribed by US physicians and provided by US pharmacies were examined using a similar methodology to the above. The results are below. These served mainly to corroborate our hypothesis that authentic Adderall samples would test negative on methamphetamine testing strips, indicating that methamphetamine-positive samples in the current study were likely counterfeit.

| Medication | FTIR Results | Amphetamine Test Strip Result | Methamphetamine Test Strip Result |
|---|---|---|---|
| Adderall XR (10mg) | Sucrose | Positive | Negative |
| Generic Amphetamine Salts-Instant Release (10mg) | Microcrystalline cellulose; DL-amphetamine sulfate | Positive | Negative |

**Resource List**

Bruker Alpha II FTIR spectrometer with platinum ATR sampling module

Dedicated testing laptop equipped with OPUS software and reference library files

Laptop to input information in the drug checking clinic survey

Charging cables for laptops and spectrometer

Ethernet cable

Nitrile gloves

KN95 masks

Disposable plastic receptacles

Razor blades

Alcohol prep pads

Kimwipes

Aluminum foil

Aluminum tray

10 mg disposable plastic micro-scoops

Disposable plastic spatulas

BTNX Rapid Response lateral flow immunoassay Benzodiazepine test strips

BTNX Rapid Response lateral flow immunoassay Fentanyl test strips

BTNX Rapid Response lateral flow immunoassay Methamphetamine test strips

BTNX Rapid Response lateral flow immunoassay Amphetamine test strips

120 mL Glass beaker

Simon’s reagents

Ceramic plate

IKA lab dancer model vortex mixer

3 mL Sterile water ampules

1.5 mL Eppendorf vials

Disposable plastic cups

DisposeRx packets

20-dram disposable plastic pill bottle

Clorox wipes

Cardboard receptacle to ensure safe transport of FTIR

Extension cord

Power strip

Lantern

6ft foldable table

2 chairs

Tape

Sharpie

Index cards

Jackery Explorer 1000 Portable Power Station

Trash bags

**Pharmacy Level Results**

| Pharmacy Number | Controlled Substances Offered in Any Form (Bottles or Single Pills) | Single Pills Sold | Counterfeit Pills Obtained | Methamphetamine Obtained | Fentanyl Obtained | Heroin Obtained |
|---|---|---|---|---|---|---|
| 1 | Y | N | N | N | N | N |
| 2 | Y | Y | Y | Y | N | N |
| 3 | Y | Y | N | N | N | N |
| 4 | Y | Y | N | N | N | N |
| 5 | N | N | N | N | N | N |
| 6 | Y | N | N | N | N | N |
| 7 | N | N | N | N | N | N |
| 8 | Y | N | N | N | N | N |
| 9 | Y | N | N | N | N | N |
| 10 | Y | N | N | N | N | N |
| 11 | Y | N | N | N | N | N |
| 12 | Y | Y | Y | Y | Y | N |
| 13 | N | N | N | N | N | N |
| 14 | Y | Y | Y | Y | N | N |
| 15 | N | N | N | N | N | N |
| 16 | Y | N | N | N | N | N |
| 17 | Y | Y | N | N | N | N |
| 18 | Y | Y | N | N | N | N |
| 19 | Y | Y | N | N | N | N |
| 20 | Y | Y | Y | Y | N | N |
| 21 | Y | N | N | N | N | N |
| 22 | N | N | N | N | N | N |
| 23 | N | N | N | N | N | N |
| 24 | Y | Y | Y | Y | N | N |
| 25 | N | N | N | N | N | N |
| 26 | N | N | N | N | N | N |
| 27 | N | N | N | N | N | N |
| 28 | N | N | N | N | N | N |
| 29 | Y | Y | N | N | N | N |
| 30 | N | N | N | N | N | N |
| 31 | Y | Y | Y | Y | Y | N |
| 32 | Y | Y | Y | Y | Y | Y |
| 33 | Y | Y | Y | N | Y | N |
| 34 | Y | Y | Y | N | Y | N |
| 35 | Y | N | N | N | N | N |
| 36 | Y | Y | N | N | N | N |
| 37 | Y | Y | Y | N | N | Y |
| 38 | Y | Y | Y | N | Y | Y |
| 39 | N | N | N | N | N | N |
| 40 | Y | Y | N | N | N | N |
